# Hypertension, Intracranial Arteriosclerosis, and Structural Brain Changes in Patients with TIA or Ischemic Stroke

**DOI:** 10.1101/2024.08.25.24312560

**Authors:** Xi Li, Bernhard P. Berghout, Gijs van Rooijen, M. Kamran Ikram, Bob Roozenbeek, Daniel Bos

## Abstract

**Background:** Hypertension is a major risk factor of structural brain changes, including atrophy and cerebral small vessel disease. Intracranial arteriosclerosis could be an underlying mechanism between hypertension and structural brain changes. This study investigated whether intracranial carotid artery calcification (ICAC), as a proxy for intracranial arteriosclerosis, explains the association between hypertension and structural brain changes in patients with TIA or ischemic stroke.

**Methods:** 968 patients (mean age 62.7 years) with TIA or ischemic stroke from a registry study who underwent non-contrast CT (NCCT) and CT-angiography (CTA) were included in this cross-sectional study. Presence and volume (mm^3^) of ICAC were assessed on CTA. Subtypes of ICAC were assessed on NCCT, where ICAC was categorized into intimal and internal elastic lamina (IEL) type calcification. Structural brain changes, indicated by atrophy, periventricular and deep white matter lesions (WML), and lacunes were assessed on NCCT. Causal mediation analysis was performed using ICAC, ICAC volume, and ICAC subtypes as mediators.

**Results:** ICAC was prevalent in 67.8% of patients, with 52.6% of them exhibiting intimal calcification, and 26.5% exhibiting IEL calcification. Atrophy, periventricular WML, deep WML, and lacunes were present in 48.1%, 56.4%, 43.0% and 17.1% of patients respectively. The presence of ICAC explained 7.1% of the association of hypertension with periventricular WML, 3.6% with deep WML, and 17.6% with lacunes. Hypertension was associated with increased atrophy through ICAC (OR: 1.02, 95% CI: 1.00-1.05). In subgroup analyses, IEL calcification partly explained the association between hypertension and periventricular WML (16.8%), and atrophy (OR: 1.12, 95% CI: 1.02-1.27). Intimal calcification did not explain any association.

**Conclusion:** ICAC partially explained the association between hypertension and atrophy, periventricular and deep WML, and lacunes. Although intimal calcification was more prevalent in ischemic stroke patients, IEL calcification takes the leading role in explaining the association between hypertension and structural brain changes.

## Introduction

Structural brain changes such as brain atrophy, white matter lesions (WML), and lacunes are common in the stroke population, and contribute significantly to post-stroke cognitive impairment^1^. Within the complex etiological framework of structural brain changes, hypertension is considered as a leading modifiable risk factor,^2^ yet the intricate pathophysiology linking hypertension with structural brain changes remains unclear.

One plausible explanation lies in arteriosclerosis of intracranial internal carotid arteries, which is the major vessel bed closest to the brain, and is prominently associated with vascular brain disease.^3,4^ Intracranial arteriosclerosis is highly prevalent among ischemic stroke patients and reported to contribute to 75% of the risk of all strokes, emphasizing its clinical importance.^5^ Intracranial carotid artery calcification (ICAC) is often used as a proxy indicator for intracranial arteriosclerosis,^6^ which is characterized by two distinct morphological subtypes: one involving the intimal layer, and the other involving the internal elastic lamina (IEL) of the artery.^7^ Intimal calcification is commonly considered as a marker of atherosclerotic plaque and related to luminal narrowing. On the other hand, thin and circular calcification in the IEL is related to arterial stiffening, affecting artery compliance and vasodilation.

Hypertension plays a pivotal role in the development of ICAC, with elevated blood pressure inducing remodeling of arterial wall structure and impairing endothelial function.^2^ Consequently, it can compromise the autoregulation of cerebral blood flow and lead to brain tissue damage.^8^ Hence, ICAC has been suggested as a mediator explaining the association between hypertension and structural brain changes. Prior research in the general population identified IEL calcification as the predominant ICAC subtype, highlighting it as the leading mechanism explaining the link between blood pressure and cerebral small vessel diseases.^9^ In contrast, ischemic stroke patients may exhibit a higher prevalence of intimal calcification as it is considered as a major risk factor of cerebral ischemia.^5^ However, no prior study has investigated the scope of the effects of these ICAC subtypes on structural brain changes in ischemic stroke patients, whereas such a study may reveal the etiological pathway of hypertension on cognitive dysfunctioning related to these structural brain changes. Considering this clinical importance of ICAC, we aimed to investigate the prevalence of the different subtypes of ICAC and to assess the magnitude of their contribution to the relation between hypertension and structural brain changes in patients with TIA and ischemic stroke.

## Methods

### Study population

We selected 1492 patients from the Erasmus Stroke Study, a cross-sectional registry study of patients with cerebrovascular diseases admitted to Erasmus MC, Rotterdam, the Netherlands from December 2005 to October 2010.^10^ Patients included in the ESS underwent clinical evaluation at presentation in either the outpatient clinic, emergency care department, or neurology ward. As routine clinical care, these patients underwent blood sampling, non-contrast CT (NCCT) and CT angiography (CTA) imaging. Patients with a TIA or ischemic stroke event with available CT and CTA scans were included in this study. Patients were excluded if data on both antihypertensive drug use and blood pressure values were not available. Patients were also excluded if the results were different on CT and CTA scans regarding the presence of ICAC, as this inconsistency may be attributed to difficulties in detecting small calcification on CT scans with thick slices or distinguishing calcification on CTA scans due to the similar density of contrast and thin calcification. This study followed the STROBE (Strengthening the Reporting of Observational Studies in Epidemiology) and AGreMA (A Guideline for Reporting Mediation Analyses) reporting guidelines. ^11,12^

### Image acquisition

Imaging was performed using a 16-slice, 64-slice, or 128-slice multidetector CT (MDCT) system (Brilliance 64, Philips Healthcare Systems, Eindhoven, Netherlands; Sensation 16, Sensation 64, Definition, Definition AS+ or Definition Flash, Siemens Medical Solutions, Erlangen, Germany) with a standardized optimized contrast-enhanced protocol. The scans ranged from the ascending aorta to the intracranial circulation (3 cm above the sella turcica). Detailed information on the scanning protocol is provided elsewhere.^10^ All MDCTA and MDCT scans were evaluated by trained readers blinded for clinical data.

### Assessment of intracranial carotid artery calcification

Calcification in the intracranial carotid artery was evaluated on axial CT angiography (CTA) images, from the start of the petrous carotid canal until the circle of Willis. Upon identification, we manually delineated closely the arterial calcification per slice. Using this region of interest, a threshold of 600 Hounsfield units (HU) on CTA scans was then applied to differentiate arterial calcification from contrast material in the lumen.^13^ Calcification volume (mm^3^) was then calculated by multiplying the number of pixels above 600HU, pixel-size, and the slice increment. The total volume of ICAC was calculated by summing up calcification volumes of both left and right intracranial carotid arteries.

A previously validated method based on CT images was applied to differentiate intimal and IEL calcification subtypes among the ICAC.^7^ This method comprises a composite score consisting of specific weighting for calcification circularity, thickness, and continuity. Both left and right carotid arteries were categorized into having predominantly intimal calcification (<7 points; i.e. thick, non-circular, and irregular), predominantly IEL calcification (≥7 points; i.e. elongated, circular, and thin), or no calcification. The intra-rater reliability of scoring ICAC subtypes was evaluated in 30 randomly-selected scans (Cohen kappa value was 0.850 for the right intracranial carotid artery, and 0.834 for the left). Based on the different combinations, we classified patients into four subtype groups. Patients with bilateral predominant intimal calcifications or intimal calcifications combined with absent contralateral calcifications were classified as intimal type ICAC. Similarly, patients with bilateral predominant IEL calcifications or IEL calcifications combined with absent contralateral calcifications were classified as IEL type ICAC. Patients with predominant intimal or IEL calcifications on one side and a contrasting contralateral subtype were classified as the mixed type. Patients without calcifications were classified as absence of ICAC.

### Assessment of structural brain changes

Brain atrophy, periventricular and deep WML, and lacunes detected on CT scans were used as imaging markers for structural brain changes. Trained researchers rated the degree of atrophy, periventricular and deep WML, and the presence of lacunes on NCCT scans. A simplified ‘Generalized Pasquier Scale’ was used to visually assess global cortical atrophy (GCA) on NCCT images.^14^ The semioval centre was first identified on axial CT images, whereafter the degree of general cerebral atrophy was rated from a 0 (not atrophic) to 3 (‘knifeblade’ atrophy) scale on that particular image, according to the original 13-region Pasquier method. The Fazekas scale^15^ was applied to assess white matter changes and it comprises two 4-point (grade 0-3) scales for assessing periventricular and deep WML respectively. Grade 0 = absent; 1 = pencil-thin lining periventricular lesions or punctate focal deep WML; 2 = smooth halo periventricular lesions or early confluence of focal WML; 3 = irregular periventricular lesions extending into the deep white matter or large confluent deep WML. Lacunes were defined as 3 to 15 mm cerebrospinal fluid (CSF)-filled cavities in the sub-cortical regions, with the same density as CSF on NCCT.^16^

### Assessment of cardiovascular risk factors

Clinical, demographic and serological measures and information regarding on cardiovascular risk factors and medication use were collected from hospital medical records at admission to the hospital. For patients with ischemic stroke, blood pressure was measured on day two to five of the hospital admission. For patients with TIA, blood pressure was measured during the diagnostic workup in the outpatient department. Hypertension was defined as the use of antihypertensive medication before the inclusion event to the hospital, or a systolic blood pressure ≥140mmHg and/or diastolic blood pressure ≥90mmHg. Hypercholesterolemia was defined as the use of cholesterol lowering drugs before admission to the hospital or serum total cholesterol ≥ 6.2mmol/L. Diabetes mellitus was defined as the use of antidiabetic medication before the inclusion event, or fasting plasma glucose level ≥7.0 mmol/L and/or a 2-hour postload glucose level ≥11.0mmol/L. Smoking status was assessed at the time of the stroke event admission and categorized into never or ever smoker. Information on the history of cardiovascular diseases (including previous TIA or ischemic stroke events, ischemic heart disease, atrial fibrillation, peripheral artery disease or other vascular diseases) was collected.

### Statistical analysis

Patient characteristics are presented as means and standard deviations for normally distributed continuous data, medians with interquartile ranges for skewed variables, or frequencies and percentages for categorical variables. Due to the right skewed distribution of ICAC volume, for patients with ICAC, we log-transformed ICAC volume.

To investigate how much of the association between hypertension and structural brain changes can be explained by the presence, the volume, and the subtypes of ICAC, we performed causal mediation analysis.^17,18^ Figure 1 illustrates the hypothetical causal model that we investigated, in which the exposure is hypertension, the outcome is structural brain changes, consisting of the four separate imaging markers, and the mediator is ICAC, defined as the presence of ICAC, log-transformed volume of ICAC, or ICAC subtypes.

**Figure 1.**
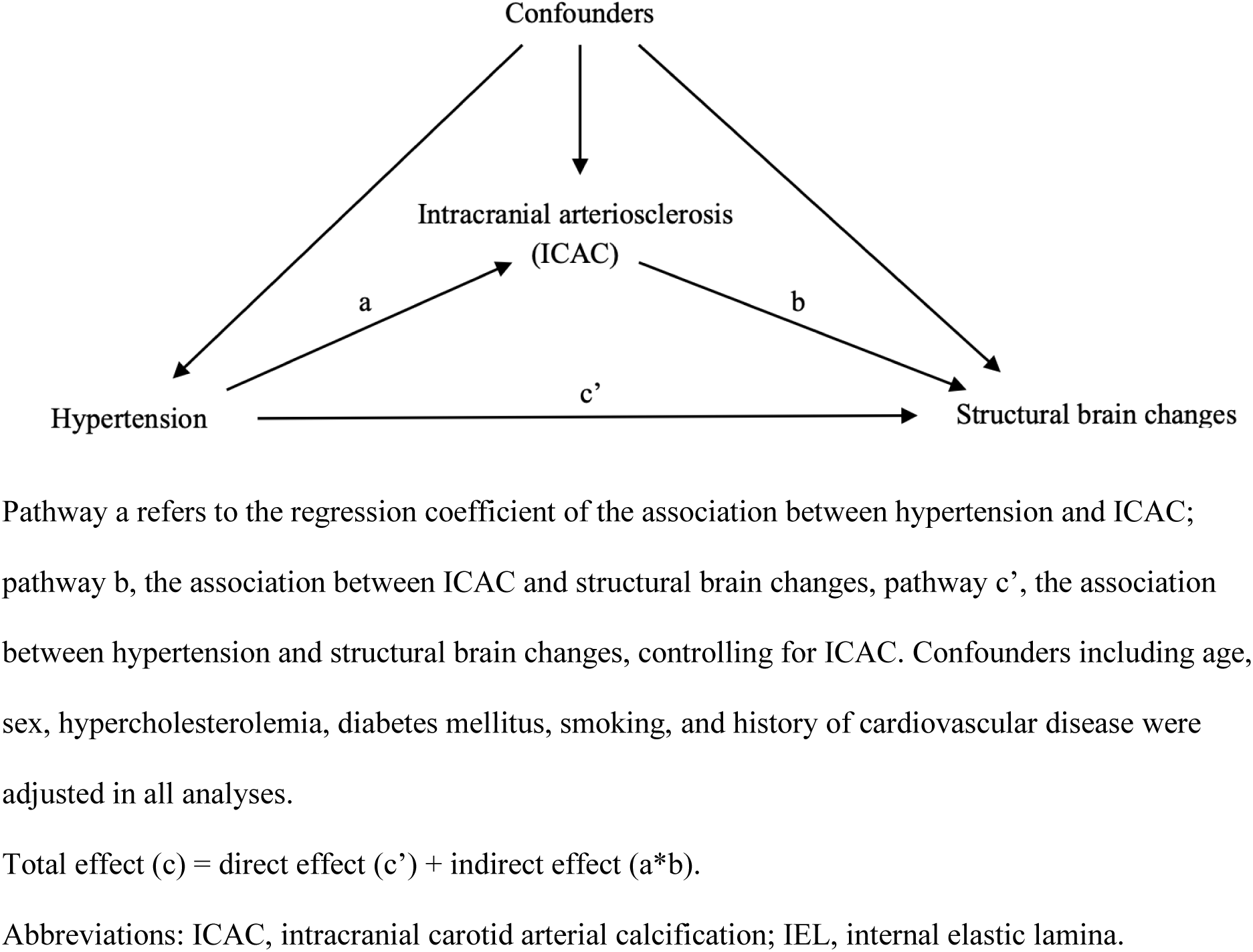
Directed acyclic graph for the association between hypertension, intracranial carotid arterial calcification (ICAC) and structural brain changes.

To perform mediation analysis, it is necessary to first test the three pathways (Figure 1): step 1, the association between hypertension and each marker of structural brain changes (total effect); step 2, the association between hypertension and ICAC (pathway a); step 3, the association between ICAC and each marker of structural brain changes (pathway b).

We analyzed the association between hypertension and the presence of ICAC with a logistic regression model. For patients with ICAC, the association between hypertension and log-transformed ICAC volume was analyzed with linear regression. We also conducted multinomial logistic regression to determine the association of hypertension with each ICAC subtype, taking absent ICAC as the reference category. Ordinal logistic regression model was used to analyze the effect of hypertension and ICAC on brain atrophy and WML. Logistic regression was fitted to analyze the association of hypertension and ICAC with the presence of lacunes. All primary analyses were performed for two models. The first model was adjusted for age and sex only. For the second model, we additionally adjusted for cardiovascular risk factors including hypercholesterolemia, diabetes mellitus, smoking, and history of cardiovascular diseases.

After all three pathways were estimated, mediation analysis was performed to estimate the direct and indirect effects based on the regression-based approach.^19^ The total effect of hypertension on structural brain changes was decomposed into two components: the indirect effect (i.e. the mediated effect) and the direct effect. Specifically, the indirect effect is the effect of hypertension on structural brain changes that can be explained by ICAC, and the direct effect is the effect that can be explained by any other factors rather than ICAC. The proportion of mediation was calculated by dividing the indirect effect by the total effect.

All analyses were conducted in the entire patient sample, and in two distinct subgroups, the first subgroup included patients with no ICAC and those with intimal type ICAC; the second subgroup comprised patients with no ICAC and those with IEL type ICAC. Mediation analyses were performed in these two subgroups separately to compare the mediated effect of intimal and IEL type calcification. Statistical analyses were conducted using R statistical software 4.3.2 (mice 3.16.0, MASS 7.3.60, CMAverse 0.1.0 R packages). Missing values in the covariates, total cholesterol level and smoking, were less than 10% and were imputed using 10-fold multiple imputation with 10 iterations. Blood pressure values were missing for 104 patients (10.7%), and these were imputed under the assumption that the data were missing at random. A sensitivity analysis was conducted by excluding patients with imputed blood pressure values to assess the impact of imputation on the results.

## Results

### Study population

A total of 968 patients were included in the analysis (Supplement Figure S1). Table 1 summarizes the patient characteristics. The mean age of the patients included in the study was 62.7±14.1years old and 45.6% of them were female. Among patients with ICAC (n = 656; 67.8%), 345 (52.6%) patients predominantly showed intimal calcification, 174 (26.5%) with IEL calcification, and 137 (20.9%) with mixed calcification. Intimal calcification was the most prevalent subtype from age 55 to 80 in the stroke population. The prevalence of IEL calcification keeps increasing with the increase of age (Figure 2). Atrophy, periventricular WML, deep WML, and lacunes were present in 48.1%, 56.4%, 43.0% and 17.1% of patients respectively.

**Figure 2.**
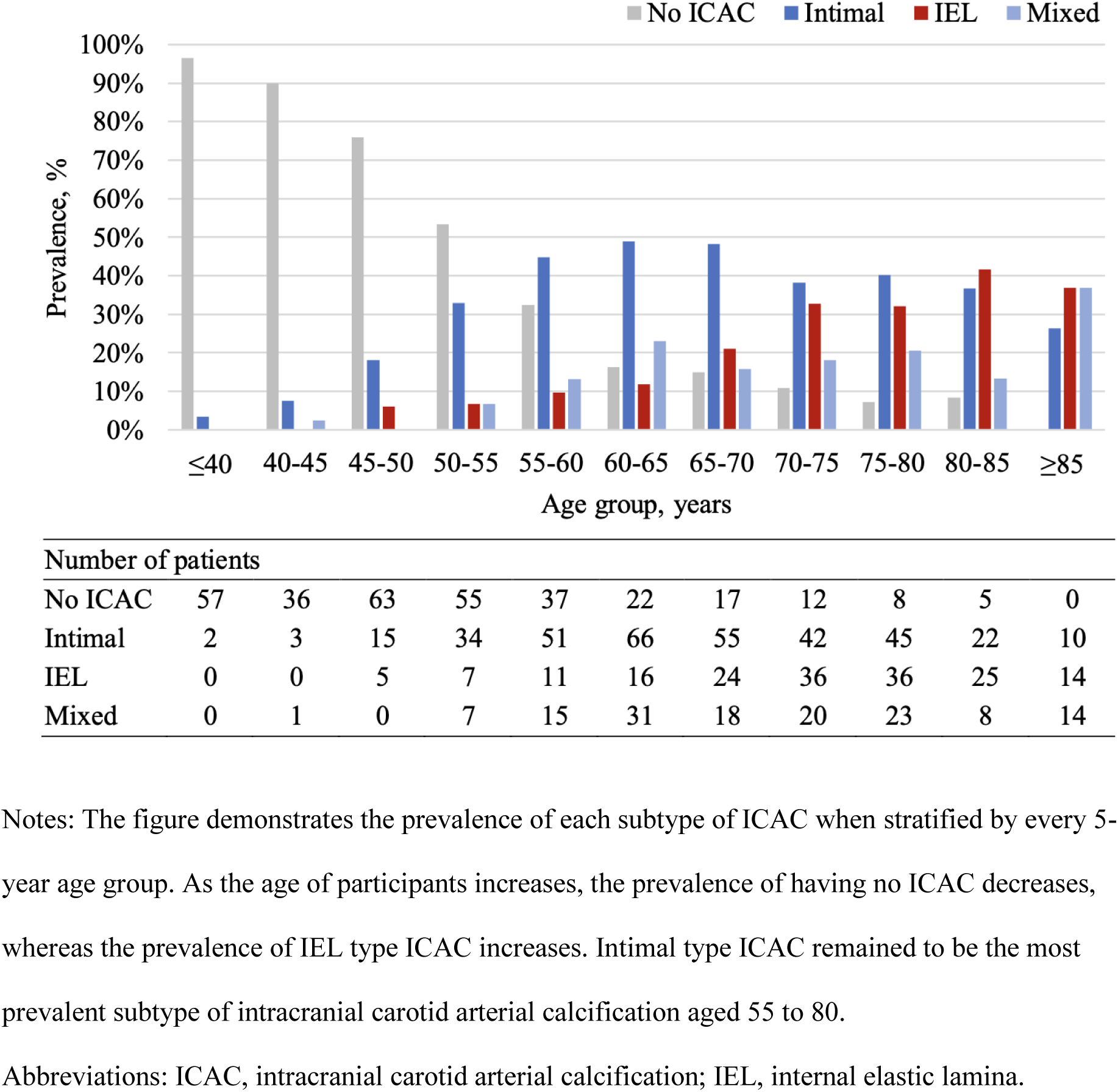
Prevalence of morphological subtypes of ICAC across every 5-year age group.

**Table 1.** Characteristics of study participants.

| Patient Characteristics | All participants (n= 968) |
| --- | --- |
| Age, years | 62.7 (14.1) |
| Women | 441 (45.6) |
| Systolic blood pressure, mmHg | 129.8 (17.7) |
| Diastolic blood pressure, mmHg | 75.5 (9.9) |
| Antihypertensive drug use | 492 (50.8) |
| Hypertension | 603 (62.3) |
| Hypercholesterolemia | 434 (44.8) |
| Diabetes Mellitus | 295 (30.5) |
| Smoking | 638 (65.9) |
| History of CVD | 460 (47.5) |
| NIHSS score | 1.0 (0.0-4.0) |
| Symptom duration < 24h (TIA) | 386 (39.9) |
| Intracranial carotid artery calcification |  |
| ICAC | 656 (67.8) |
| intimal type calcification | 345 (52.6) |
| IEL type calcification | 174 (26.5) |
| mixed type calcification | 137 (20.9) |
| Calcification volume, mm<sup>3</sup> | 23.1 (5.1- 69.5) |
| Structural brain changes |  |
| Atrophy | 466 (48.1) |
| Periventricular white matter lesions | 546 (56.4) |
| Deep white matter lesions | 416 (43.0) |
| Lacunae | 166 (17.1) |

Notes: Continuous variables were presented as mean ( ± SD) or median (interquartile range).
Categorical variables were shown as numbers of patients and frequencies (%).
Abbreviations: ICAC, intracranial carotid artery calcification. IEL, internal elastic lamina.

### Association between hypertension, ICAC and structural brain changes

Hypertension was associated with the prevalence of ICAC (adjusted Odds Ratio (aOR): 1.63, 95% CI: 1.11-2.38), in particular with IEL calcification (aOR: 2.49, 95% CI: 1.45-4.28) (Table 2). Similarly, hypertension was associated with more severe periventricular WML (adjusted common Odds Ratio (acOR): 1.50, 95% CI: 1.13-2.00) and deep WML (acOR: 1.82, 95%CI: 1.34-2.47) (Table 3). The presence of ICAC was independently associated with more severe atrophy (acOR: 2.15, 95% CI: 1.44-3.20), periventricular WML (acOR: 1.84, 95% CI: 1.30-2.62), deep WML (acOR: 1.49, 95% CI 1.02-2.18), and the presence of lacunes (aOR: 1.70, 95% CI: 1.01-2.88). With absent ICAC being the reference group, IEL calcification has a more pronounced association with atrophy (acOR: 2.67, 95% CI 1.62-4.40), periventricular WML (acOR: 1.93, 95% CI 1.24-3.01) and deep WML (acOR: 1.69, 95% CI 1.06-2.70) than intimal calcification (Table 3). For lacunes, the only significant association was with mixed calcification (aOR: 2.18, 95% CI 1.15-4.13).

**Table 2.**
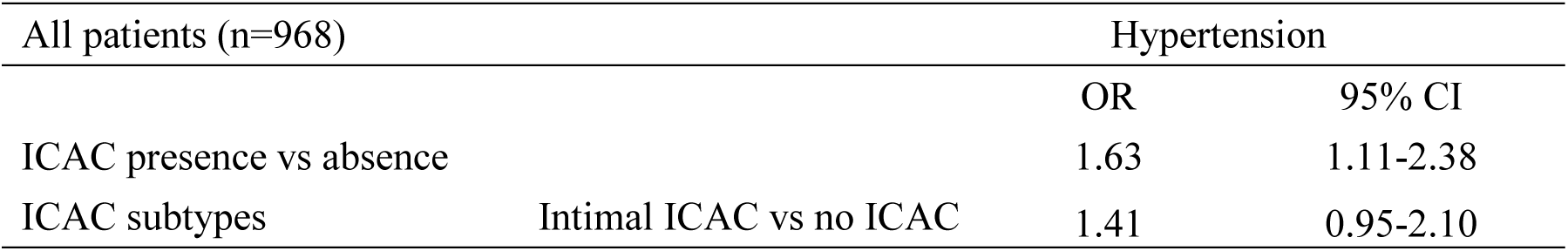
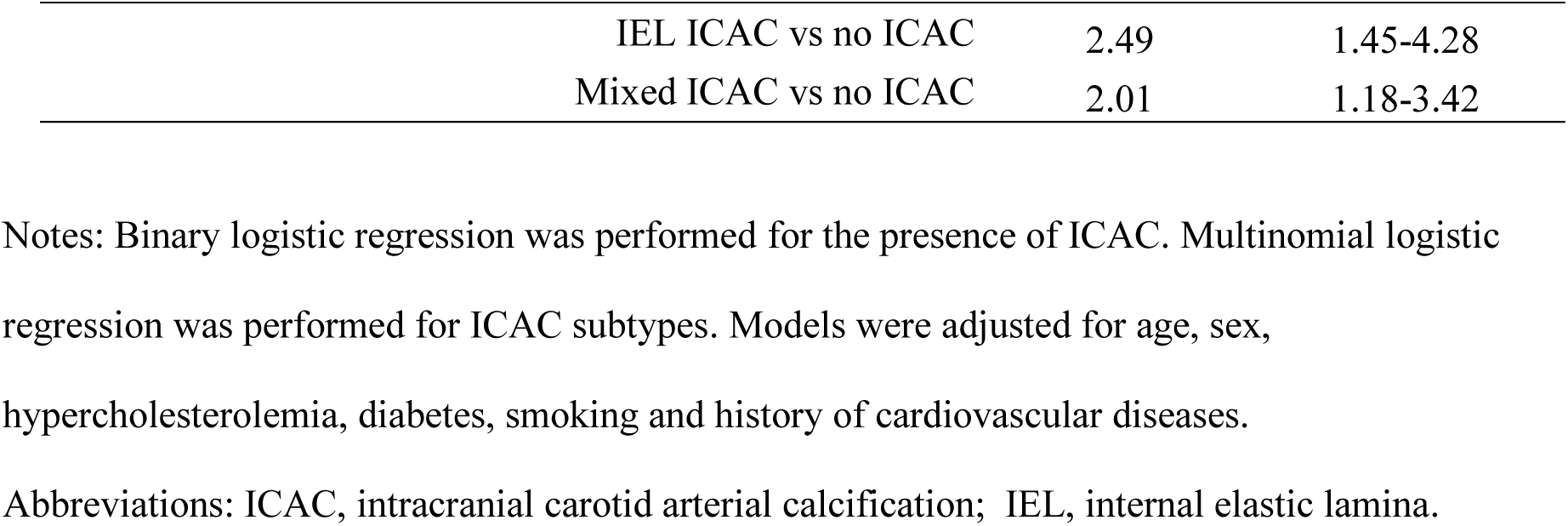
Association of hypertension with the presence and subtypes of intracranial carotid arterial calcification (ICAC)

| All patients (n=968) |  | Hypertension |  |
| --- | --- | --- | --- |
|  |  | OR | 95% CI |
| ICAC presence vs absence |  | 1.63 | 1.11-2.38 |
| ICAC subtypes | Intimal ICAC vs no ICAC | 1.41 | 0.95-2.10 |

|  |  |  |
| --- | --- | --- |
| IEL ICAC vs no ICAC | 2.49 | 1.45-4.28 |
| Mixed ICAC vs no ICAC | 2.01 | 1.18-3.42 |

Notes: Binary logistic regression was performed for the presence of ICAC. Multinomial logistic regression was performed for ICAC subtypes. Models were adjusted for age, sex, hypercholesterolemia, diabetes, smoking and history of cardiovascular diseases.
Abbreviations: ICAC, intracranial carotid arterial calcification; IEL, internal elastic lamina.

**Table 3.**
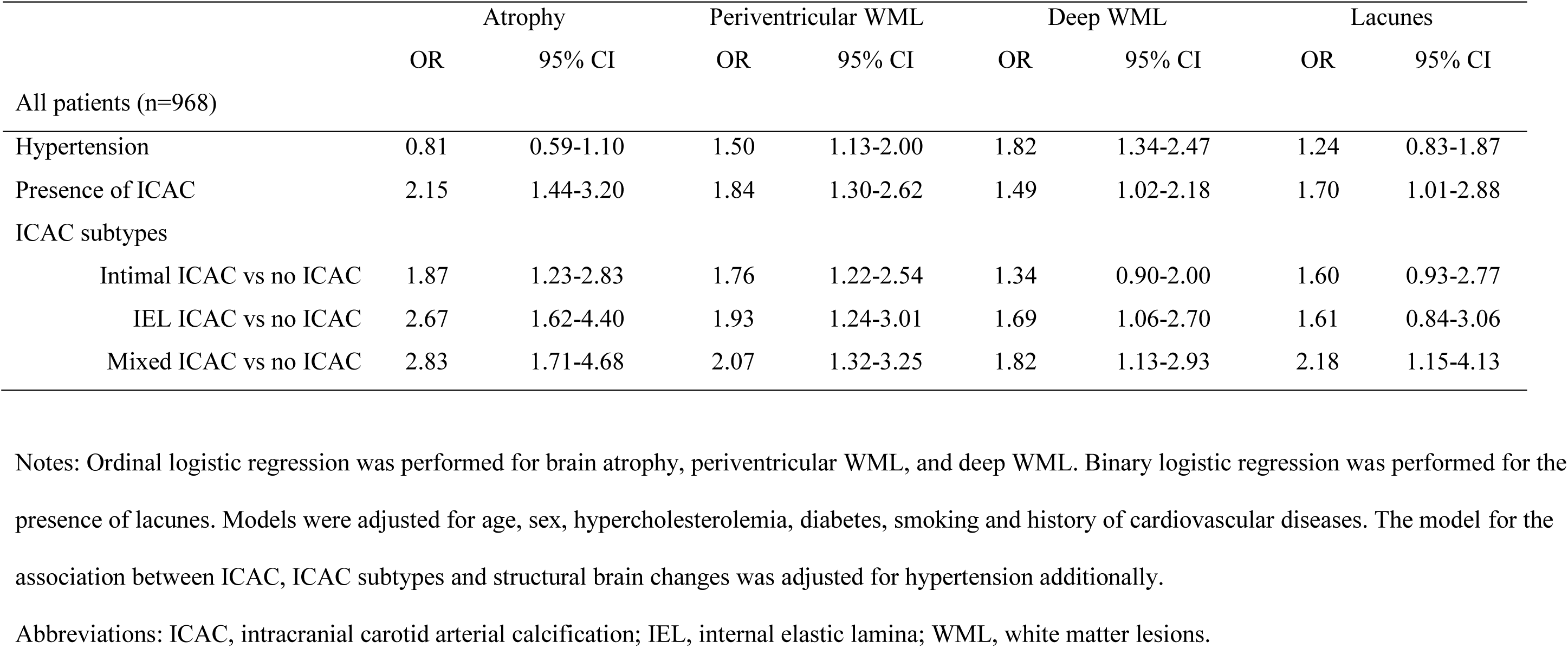
Association of hypertension and intracranial carotid arterial calcification (ICAC) with structural brain changes.

### ICAC as a mediator between hypertension and structural brain changes

In the mediation analyses, the presence of ICAC explained 7.1% of the association of hypertension with periventricular WML, 3.6% of the association with deep WML, and 17.6% with the presence of lacunes (Table 4). Hypertension was associated with more severe brain atrophy through the mediation of ICAC (acOR: 1.02, 95% CI: 1.00-1.05), while the direct effect of hypertension on brain atrophy was towards the opposite direction (acOR: 0.82, 95% CI 0.64-1.04).

**Table 4.**
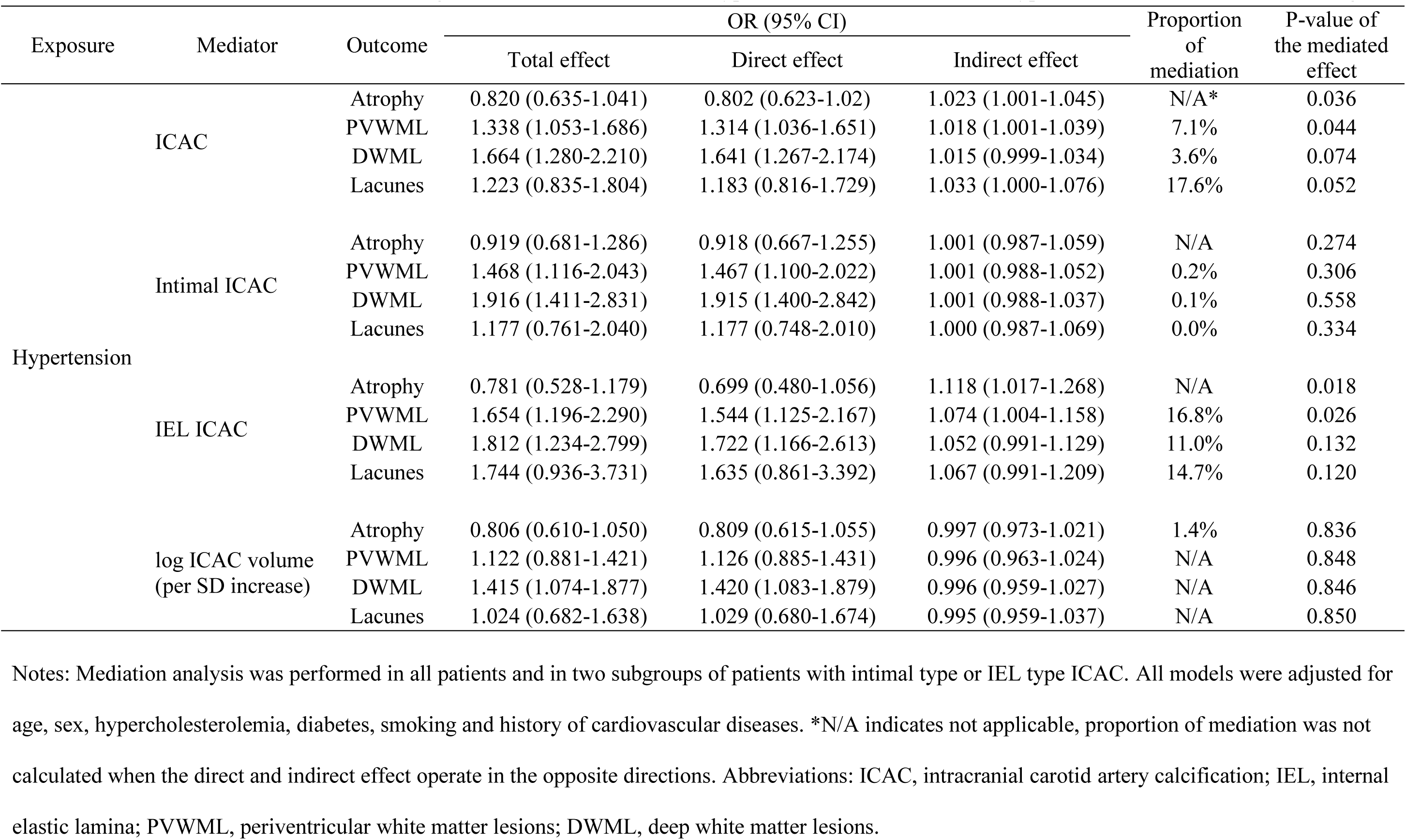
Estimated mediation effect of ICAC, log-ICAC volume, and ICAC subtypes in the association between hypertension and structural brain changes.

| Exposure | Mediator | Outcome | OR (95% CI) |  |  | Proportion of mediation | P-value of the mediated effect |
| --- | --- | --- | --- | --- | --- | --- | --- |
|  |  |  | Total effect | Direct effect | Indirect effect |  |  |
| Hypertension | ICAC | Atrophy | 0.820 (0.635-1.041) | 0.802 (0.623-1.02) | 1.023 (1.001-1.045) | N/A* | 0.036 |
|  |  | PVWML | 1.338 (1.053-1.686) | 1.314 (1.036-1.651) | 1.018 (1.001-1.039) | 7.1% | 0.044 |
|  |  | DWML | 1.664 (1.280-2.210) | 1.641 (1.267-2.174) | 1.015 (0.999-1.034) | 3.6% | 0.074 |
|  |  | Lacunes | 1.223 (0.835-1.804) | 1.183 (0.816-1.729) | 1.033 (1.000-1.076) | 17.6% | 0.052 |
|  | Intimal ICAC | Atrophy | 0.919 (0.681-1.286) | 0.918 (0.667-1.255) | 1.001 (0.987-1.059) | N/A | 0.274 |
|  |  | PVWML | 1.468 (1.116-2.043) | 1.467 (1.100-2.022) | 1.001 (0.988-1.052) | 0.2% | 0.306 |
|  |  | DWML | 1.916 (1.411-2.831) | 1.915 (1.400-2.842) | 1.001 (0.988-1.037) | 0.1% | 0.558 |
|  |  | Lacunes | 1.177 (0.761-2.040) | 1.177 (0.748-2.010) | 1.000 (0.987-1.069) | 0.0% | 0.334 |
|  | IEL ICAC | Atrophy | 0.781 (0.528-1.179) | 0.699 (0.480-1.056) | 1.118 (1.017-1.268) | N/A | 0.018 |
|  |  | PVWML | 1.654 (1.196-2.290) | 1.544 (1.125-2.167) | 1.074 (1.004-1.158) | 16.8% | 0.026 |
|  |  | DWML | 1.812 (1.234-2.799) | 1.722 (1.166-2.613) | 1.052 (0.991-1.129) | 11.0% | 0.132 |
|  |  | Lacunes | 1.744 (0.936-3.731) | 1.635 (0.861-3.392) | 1.067 (0.991-1.209) | 14.7% | 0.120 |
|  | log ICAC volume (per SD increase) | Atrophy | 0.806 (0.610-1.050) | 0.809 (0.615-1.055) | 0.997 (0.973-1.021) | 1.4% | 0.836 |
|  |  | PVWML | 1.122 (0.881-1.421) | 1.126 (0.885-1.431) | 0.996 (0.963-1.024) | N/A | 0.848 |
|  |  | DWML | 1.415 (1.074-1.877) | 1.420 (1.083-1.879) | 0.996 (0.959-1.027) | N/A | 0.846 |
|  |  | Lacunes | 1.024 (0.682-1.638) | 1.029 (0.680-1.674) | 0.995 (0.959-1.037) | N/A | 0.850 |

Notes: Mediation analysis was performed in all patients and in two subgroups of patients with intimal type or IEL type ICAC. All models were adjusted for age, sex, hypercholesterolemia, diabetes, smoking and history of cardiovascular diseases. \*N/A indicates not applicable, proportion of mediation was not calculated when the direct and indirect effect operate in the opposite directions. Abbreviations: ICAC, intracranial carotid artery calcification; IEL, internal elastic lamina; PVWML, periventricular white matter lesions; DWML, deep white matter lesions.

For mediation analyses performed in the two subgroups of patients with no ICAC + patients with intimal type ICAC(n = 657) and patients with no ICAC + patients with IEL type ICAC (n = 486), IEL calcification explained 16.8% of the association between hypertension and periventricular WML. The mediated effect of IEL calcification on brain atrophy was significant (acOR: 1.12, 95% CI: 1.02-1.27) but on the opposite direction of the direct effect of hypertension on brain atrophy (acOR: 0.78, 95% CI: 0.53-1.18). No significant mediated effect of intimal calcification was observed. We also tested the interaction between hypertension and ICAC, but we didn’t find significant interaction effect, thus interaction terms were left out in further mediation analyses.

In the sensitivity analysis, we excluded 104 patients with missing values on blood pressure, and ended up with 864 patients for the mediation analysis. The mediation effects found were similar (Supplement Table S3).

## Discussion

In this cross-sectional study of patients with TIA or ischemic stroke, the presence of ICAC was observed in two-thirds of the patients. Our mediation analyses results revealed that the presence of ICAC partially explained the effect of hypertension on brain atrophy, periventricular WML, deep WML, and lacunes. Despite the higher prevalence of intimal calcification in stroke patients, the mediated effect was mainly driven by IEL calcification.

### Prevalence of intimal and IEL calcification in patients with ischemic stroke or TIA

This study found that intimal calcification was the most prevalent type of intracranial arteriosclerosis in patients with TIA or ischemic stroke, which is consistent with the recognized involvement of atherosclerosis in the pathophysiology of cerebral ischemia.^5^ However, a previous study reported IEL calcification to be the most prevalent type in ischemic stroke patients.^20^ This could be explained by the younger age of our study participants (mean age: 62 years old), as IEL calcification is strongly associated with older age. Stratifying the prevalence by every 5-year age group revealed an increasing predominance of the IEL subtype with advancing age, which aligns with previous research.^21^ These findings highlight that calcification along the IEL of intracranial arteries is an age-related process.

### Association between hypertension, ICAC and structural brain changes

The effect of hypertension and ICAC varies between different markers of structural brain changes. The opposite direction of direct and indirect effect on brain atrophy indicates that while higher blood pressure may serve as a compensatory mechanism for inadequate cerebral blood flow, ^22,23^ the presence of ICAC counteracts with this protective effect and leads to increased atrophy, especially for IEL calcification. For lacunes, although no significant direct effect of hypertension was found, the effect through mediation of ICAC was significant. This is consistent with the pathophysiological perspective, where hypertension contributes to lacune formation by inducing changes in small arteries, and increasing their susceptibility to occlusion.^24^ Compared to atrophy and lacunes, the direct effect of hypertension on WML was substantially larger and remained to be significant. This implies that while hypertension is a strong risk factor of WML, the effect primarily operates through other pathways than ICAC, such as inflammation and oxidative stress^2^, and increased blood-brain barrier permeability.^25^

### Mediated effect through intimal and IEL calcification

The mediation analyses revealed the small but significant contribution of ICAC to the effect of hypertension on structural brain changes. Comparing the mediated effect of the two subtypes of ICAC, IEL calcification explained a larger proportion of the effect of hypertension on structural brain changes. This was within our expectation as more pronounced association was observed between hypertension and IEL calcification (pathway a), and between IEL calcification and structural brain changes (pathway b). A larger mediated effect of IEL calcification was also observed in a prior study conducted in the general population (age 68.0±5.7 years) where IEL calcification was the most common form of ICAC.^9^ This implies that the prevalence of ICAC subtypes may change with age and according to whether people suffered from stroke, but IEL calcification remains the leading mechanism explaining the association between hypertension and structural brain changes. The potential mechanism involves IEL calcification as an indicator of arterial aging and stiffness, contributing to impaired cerebral autoregulation and the progression of neurodegenerative processes by facilitating excessive pulsatile pressure in the cerebral microcirculation.

### Strengths and limitations

This study used causal mediation analysis to explain the underlying mechanism of how hypertension is related to structural brain changes in patients with TIA or ischemic stroke, where the prevalence of ICAC and structural brain changes are different compared to the general population.

However, this study also contains several important limitations. First, with the cross-sectional design of this study, we cannot draw a causal conclusion on the effect of hypertension on ICAC. Nevertheless, the biological plausibility of hypertension leading to arteriosclerosis and subsequent brain tissue damage supports the temporal ordering of variables. Future longitudinal studies incorporating detailed blood pressure measurements over time would provide valuable insights. Second, the CT scans we used to classify ICAC subtypes were 3 to 4.8mm thick, which increased the possibility of missing small calcifications or misclassifying subtypes of calcification due to the difficulty of assessing continuity with thick slices. Lastly, for a mediation analysis with multinomial mediator, the current mediation R package does not support calculating separate mediation effects for each category of the mediator. Therefore, we chose to compare the mediation effect of two ICAC subtypes in two subgroups of patients. However, it aligns with our research aim of comparing the mediating roles of two ICAC subtypes. The more pronounced association observed between IEL calcification and hypertension, along with structural brain changes also supports our finding of the significant mediated effect of IEL calcification.

## Conclusion

This study showed that ICAC partially explained the association between hypertension and brain atrophy, periventricular and deep WML, and lacunes. Although a higher prevalence of intimal calcification was found in patients with ischemic stroke or TIA, the mediated effect was mainly driven by IEL type calcification.

## Data Availability

The data underlying this article cannot be shared publicly due to the privacy of participants in this study. However, anonymized data will be shared on reasonable request to the corresponding author.

## Acknowledgments

None.

## Informed consent and ethical approval

Written informed consent for participation in the study was obtained from all patients and the study was approved by the institutional ethics committee (Erasmus MC MERC, approval number MEC-2005-345).

